# Automated GenePy Gene-Burden Computation via a Reproducible Nextflow Workflow Integrated with the Genomics England (GEL) Lifebit Platform

**DOI:** 10.64898/2026.05.22.26353863

**Authors:** Iman Nazari, Guo Cheng, James Ashton, Sarah Ennis

## Abstract

Interpretation of rare-disease genomes remains constrained by variant-centric analytical frameworks that insufficiently capture the cumulative impact of multiple variants within a gene. GenePy provides an individual-level, gene-based burden metric that integrates variant consequence, allele frequency, and zygosity into a unified quantitative score, enabling a transition from discrete variant annotation to aggregated gene-level interpretation. In the context of Genomics England, this formulation supports a panel-agnostic, genotype-to-phenotype diagnostic strategy for unresolved monogenic disorders by prioritising genes with elevated mutational burden per individual.

Here, we present a fully automated, containerised GenePy workflow deployed through Nextflow and integrated within the Genomics England (GEL) Research Environment via the Lifebit CloudOS platform. This implementation provides scalable, secure, and governance-compliant computation of gene-level burden scores across population-scale cohorts. The workflow harmonises variant annotation, quality control, and chunked data aggregation within modular, reproducible processes designed for high-throughput execution on cloud-native infrastructure. By enabling robust, portable, and auditable gene-level scoring across large rare-disease sequencing datasets, this framework enhances analytical resolution and supports downstream statistical prioritisation, integrative phenotype matching, and hypothesis generation within genotype-to-phenotype diagnostic workflows.

## 1 Introduction

GenePy [1] is a computational framework that converts next-generation sequencing (NGS) data into individual-level gene-burden scores by integrating key variant attributes, including predicted functional consequence, allele frequency, and zygosity, into a unified metric. This formulation shifts the interpretive focus from isolated variant calls towards a more biologically coherent quantification of cumulative gene perturbation. Such gene-level representation is particularly advantageous for investigating complex diseases, in which pathogenicity typically reflects the aggregate contribution of multiple moderate-impact variants rather than singular high-effect alleles. Initial applications of GenePy have demonstrated its capacity to enhance detection of gene–disease associations, support pathway-level analyses, and integrate seamlessly with multiple deleteriousness metrics [2]. Accordingly, GenePy holds great potential for gene discovery by enabling systematic prioritisation of candidate genes for downstream validation.

The Genomics England (GEL) Research Environment [3] provides a unique platform for deploying such frameworks because of its secure, accredited architecture and access to the National Genomic Research Library, which includes large-scale whole-genome and whole-exome sequencing data linked to phenotype-rich clinical records. The strict governance model of the GEL environment, which restricts identifiable data to a secure workspace while supporting reproducible computation, necessitates the use of portable, containerised workflows that maintain consistent behaviour across heterogeneous computing environments.

Nextflow [4] offers an ideal solution for orchestrating GenePy, enabling modular, version-controlled workflow design with explicit management of dependencies and parallelisation. By packaging all components in Docker [5] or Singularity [6] containers, the workflow achieves complete reproducibility, transparent provenance, and seamless deployment across cloud, HPC, and restricted-access environments. When executed through the Lifebit CloudOS platform [7], which leverages Amazon Web Services (AWS) [8], the pipeline benefits from elastic scaling, managed scheduling, standardised metadata capture, and full compliance with GEL data-governance requirements.

By integrating GenePy within a cloud-native, containerised Nextflow framework, this work provides a scalable, reproducible, and governance-compliant solution for computing gene-level perturbation scores across population-scale genomic datasets, enabling deeper interrogation of complex trait architecture and reproducible integration with statistical and machine-learning analyses (Figure 1). The full workflow, including source code, container definitions, and configuration files, is openly available at the GitHub repository `UoS-HGIG/Genepy_GEL_V2`.

**Figure 1.**
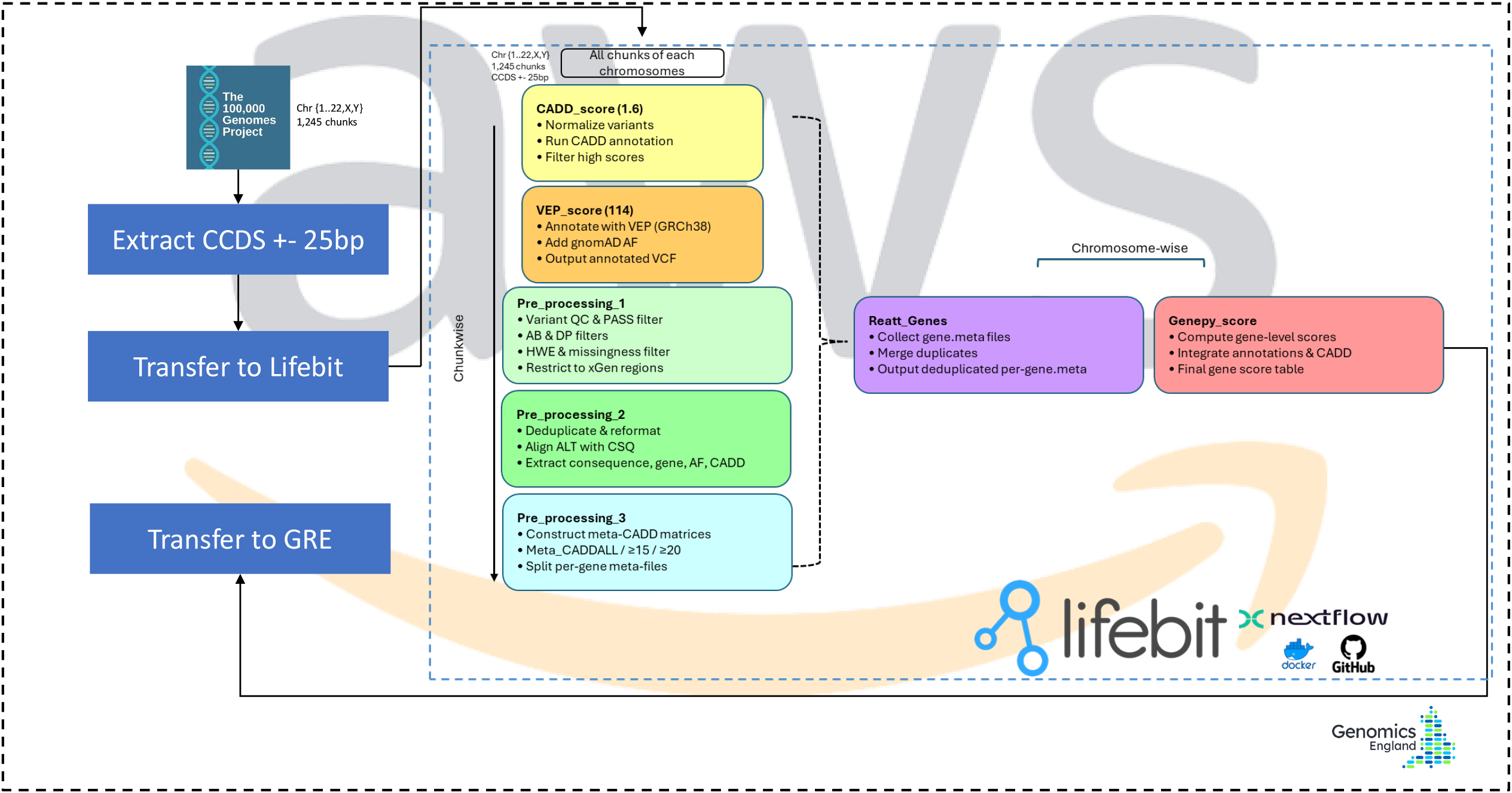
A scalable Nextflow-orchestrated workflow integrating Lifebit CloudOS and GEL infrastructure to compute GenePy gene-level pathogenicity scores, beginning with CCDS ± 25 bp extraction from 100kGP data and progressing through CADD and VEP annotation, multi-stage variant preprocessing, gene-level re-aggregation, and final score generation.

## 2 Methods

### 2.1 Data sources and processing environment

All analyses were performed within the Genomics England Research Environment using aggregated germline whole-genome sequencing (WGS) data from approximately 78,000 participants in the 100,000 Genomes Project (AGG2 release). Raw input variant data were provided as per-chromosome VCF chunks (approximately 80–200 GB per chunk) and were integrated with participant-level metadata available within the secure environment. To reduce computational and storage burden while retaining regions of primary clinical interpretability, variants were subset to CCDS coding regions plus ±25 bp flanking sequence. This extraction step was executed inside the Genomics England Research Environment, producing intermediate per-chromosome files of approximately 1–6 GB on average.

Computation was then conducted through the Lifebit CloudOS platform, which provides AWS-based elastic compute resources under Genomics England governance controls, after mounting the extracted CCDS ±25 bp dataset from the Genomics England environment into Lifebit for workflow execution. Pipeline outputs were formatted as a user-friendly matrix of gene-level pathogenicity scores per participant, enabling computationally efficient downstream analyses compared with repeated access to full WGS-scale files. This design emphasises scalability through cloud execution and automation while maintaining data governance appropriate for sensitive large-scale genomic data, transforming multi-terabyte WGS inputs into a compact analysis-ready representation.

### 2.2 Workflow design and orchestration

The GenePy workflow was implemented using Nextflow DSL2. The design comprises modular, containerised processes for variant preprocessing, annotation, harmonisation, and gene-level scoring. Workflow modules were version-controlled and executed within fixed Docker/Singularity images to ensure deterministic behaviour. Nextflow’s execution engine handled resource allocation, parallelisation of genomic chunks, and provenance capture, including command logs, container digests, and input checksums.

### 2.3 Variant normalisation and multisource annotation

Per-chromosome VCFs were partitioned into genomic windows and processed in parallel. Variants were normalised to a canonical representation and annotated using Ensembl VEP (v114) [9] with GRCh38 gene models and gnomAD [10] population allele frequencies. Predicted deleteriousness was incorporated using CADD v1.6 [11].

### 2.4 Quality control and harmonisation

A multi-criteria quality-control framework was applied [12, 13], including genotype filtering based on depth and allele balance, removal of loci failing caller-specific filters, Hardy–Weinberg equilibrium assessment [14], and missingness evaluation. Harmonisation steps resolved multiallelic representation, ensured one-to-one alignment between sequence variants and VEP annotations, and merged overlapping chunk boundaries into unified variant records. To account for sex-specific ploidy on the X chromosome, genotypes were normalised in a karyotype-aware manner so that downstream analyses could treat chrX genotypes consistently across individuals. In particular, haploid calls in males on chrX were recoded to an equivalent diploid representation to preserve the intended allele dosage and zygosity interpretation for gene-level burden scoring.

### 2.5 Gene-level data structuring

Variant annotations were aggregated into compact gene-level metadata files encoding coordinates, allele states, consequences, deleteriousness scores, allele frequencies, and QC flags. A genome-wide re-aggregation module merged per-chunk outputs into a single canonical record per gene, harmonising identifiers across Ensembl and HGNC nomenclature.

### 2.6 GenePy scoring

Gene-level perturbation scores were generated using the GenePy scoring module, which rescales CADD scores, integrates allele frequencies, encodes genotype states, and computes aggregated severity-weighted burden measures for each individual. The resulting gene × sample score matrix provides a quantitative substrate for statistical association testing, enrichment analysis, and machine-learning applications.

### 2.7 Execution summary and outputs

The workflow was executed in a chunk-wise parallelised manner on Lifebit CloudOS (AWS backend), a managed cloud platform optimised for scalable Nextflow execution, with automatic retries governed by Nextflow’s error-handling framework. The primary output is a comprehensive gene-score table (GenePy matrix); optional outputs include (1) annotated, quality-controlled VCFs for each chunk and (2) per-gene `.gene.meta` files. All generated artefacts were tracked via checksums and are fully reproducible from the committed workflow configuration and associated Docker images. AWS instance types used for execution are detailed in Table 1.

**Table 1.**
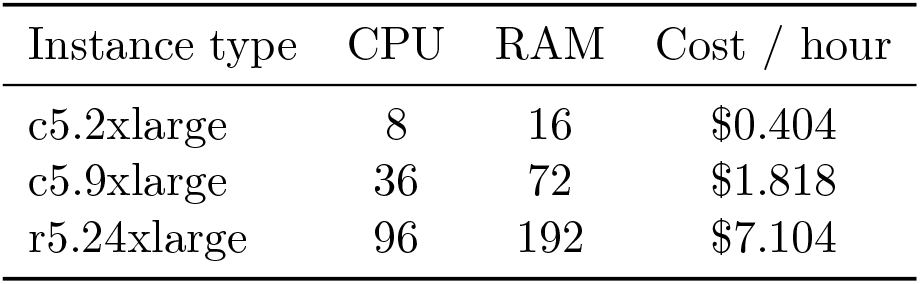
AWS instance types used in this workflow.

## 3 Results

### 3.1 GenePy score analysis reveals gene-level pathogenic burden in a panel-based rare-disease cohort

To assess the aggregate pathogenic burden across our cohort, we applied GenePy scoring, a gene-level pathogenicity metric that integrates variant-level deleteriousness predictions, population allele frequencies, and individual zygosity, to approximately 24,000 unique genes in the human reference genome across 78,000 individuals. GenePy transforms variant-level interpretation into an intuitive per-gene, per-individual score, enabling systematic comparison of genetic burden both across individuals and between case and control populations.

To examine gene-specific pathogenic-variant distributions across the cohort, we visualised GenePy density plots for three genes representing distinct disease mechanisms (Figure 2). REEP6 showed a sharp peak near zero with minimal right-tail distribution, suggesting extremely low population-level pathogenic burden and high constraint. In contrast, PRPF3 displayed a broader distribution with a secondary mode, reflecting a modest subset of individuals carrying rare deleterious variants. SERPINF1 exhibited the most complex distribution, with multiple density peaks indicating heterogeneous variant burden across the cohort, potentially reflecting both founder effects and recurrent pathogenic alleles in specific subpopulations. These density profiles provide a cohort-wide perspective on the penetrance and frequency of pathogenic variation within clinically actionable genes, facilitating prioritisation for downstream functional validation and gene–phenotype association studies.

**Figure 2.**
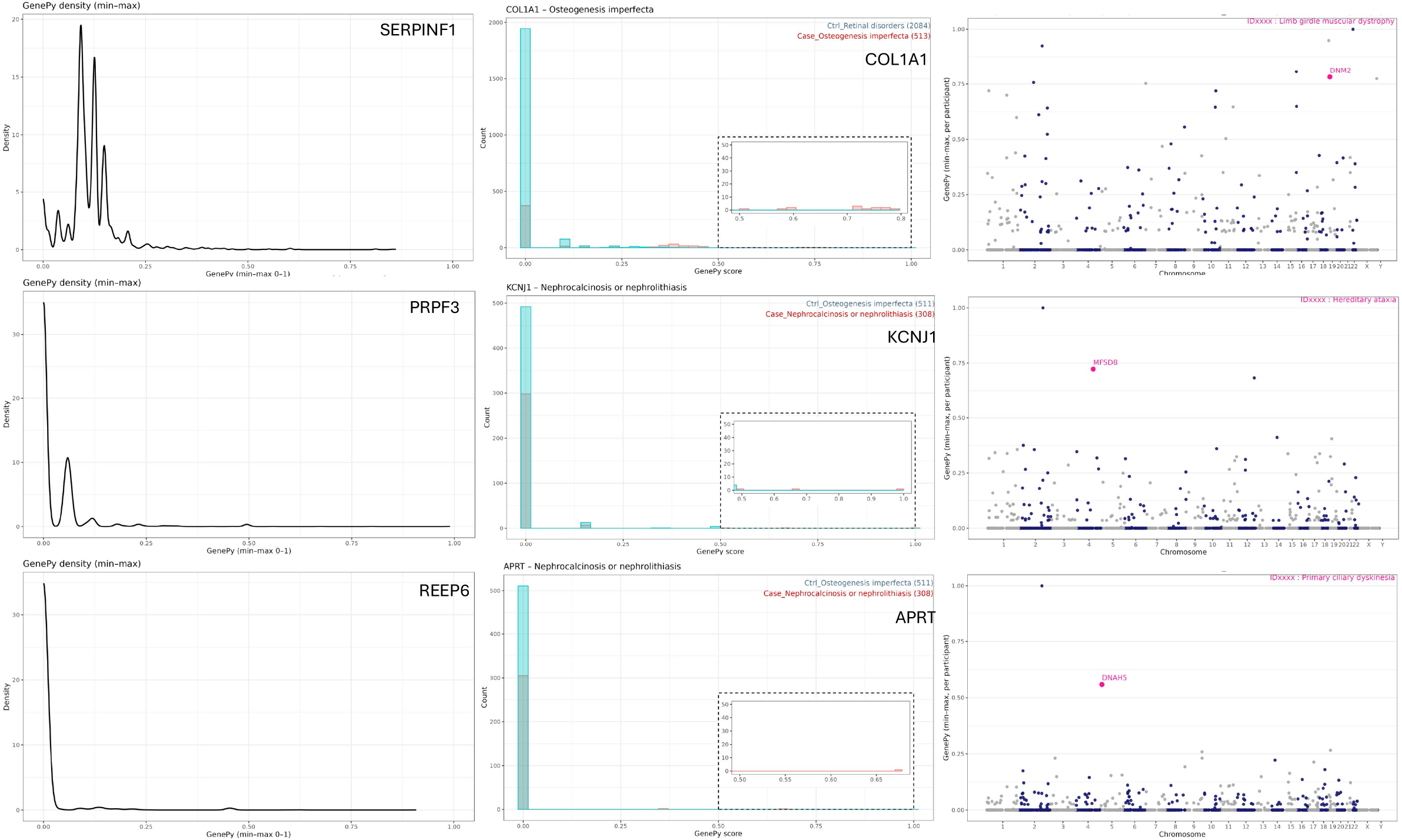
GenePy score distributions and genome-wide burden profiles. **Left:** Cohort-wide density plots for REEP6, PRPF3, and SERPINF1. **Middle:** Case–control GenePy score distributions for APRT, KCNJ1, and COL1A1. **Right:** Manhattan-style genome-wide GenePy plots for three representative patients, highlighting elevated scores in phenotype-relevant genes.

To validate the discriminatory power of GenePy in a phenotype-specific context, we compared the distribution of GenePy scores for three diagnostically relevant genes (APRT, COL1A1, and KCNJ1) between cases presenting with the corresponding phenotypes (nephrocalcinosis/nephrolithiasis for APRT and KCNJ1; osteogenesis imperfecta for COL1A1) and control individuals from the broader cohort without these diagnoses (Figure 2). All three genes demonstrated a striking bimodal distribution, with the vast majority of both case and control individuals clustering at GenePy scores near zero, reflecting the rarity of pathogenic variants in the general population. However, cases exhibited a distinct secondary peak at higher GenePy scores (inset panels), representing individuals harbouring deleterious compound-heterozygous or homozygous variants.

The genome-wide Manhattan-style plots in the rightmost column of Figure 2 illustrate the distribution of GenePy scores across all chromosomes for three representative patients, revealing elevated pathogenic scores in genes previously implicated in their respective phenotypes. Notably, DNAH5 (associated with primary ciliary dyskinesia), DNAH8 (hereditary ataxia), and DNM2 (limb-girdle muscular dystrophy) exhibited markedly elevated GenePy scores in affected individuals compared with the baseline genomic background, consistent with their clinical diagnoses obtained through structured phenotypic questionnaires.

## 4 Discussion

The implementation of GenePy within the Genomics England Research Environment enabled robust, scalable computation of gene-level burden metrics across whole-genome sequencing datasets. However, financial constraints associated with cloud-based computation necessitated a targeted analytical strategy for the AGG2 application reported here, such that cost limitations related to AWS compute utilisation precluded comprehensive genome-wide processing of all annotated loci within the available funding. Accordingly, as described in the Methods, analyses were restricted to CCDS regions with an additional ±25 bp flanking window, an interval-focused design that captures canonical coding bases and immediate splice-adjacent sites while maintaining computational affordability in the cloud-governed Lifebit execution environment.

While narrower in scope than whole-genome interrogation, this strategy retained sensitivity to variants with established functional relevance and substantially reduced computational overhead. The CCDS ±25 bp targeting should therefore be considered a configurable implementation choice rather than a fixed limitation of the pipeline. In future releases (for example, AGG3), the workflow could be extended to broader, gene-centric intervals, including whole gene bodies (coding and non-coding sequence) and expanded flanking regions spanning promoter through 5′ UTR to 3′ UTR, where inclusion of regulatory context (for example, promoters and enhancers) is desirable. Beyond single-locus intervals, the same scoring paradigm could be extended to digenic or oligogenic models by aggregating gene-level scores across biologically or phenotypically related gene sets to prioritise multi-gene hypotheses for follow-up.

Despite the restricted genomic window, this framework provided meaningful insights into the broader landscape of genetic perturbation. GenePy inherently integrates distributed variant effects into a unified, gene-level measure, enabling quantification of cumulative pathogenic burden even when variant contributions arise from modest-effect alleles. Importantly, the use of a gene-centric severity score allows signals originating from non-coding yet functionally relevant bases, such as splice-region variants, regulatory motifs adjacent to exons, or elements influencing transcript stability, to be partially captured within the defined CCDS ±25 bp interval. While the window does not encompass distal enhancers, promoters, or long-range regulatory architecture, the scoring framework nonetheless improves interpretability of near-coding variation, which frequently contributes to disease mechanisms but can be overlooked in variant-centric analyses. Because GenePy is driven by variant annotations and deleteriousness metrics, its performance in near-coding and other regulatory-proximal regions should improve as these annotations and predictors improve.

More broadly, the successful deployment of this workflow demonstrates how cloud-native, containerised pipelines can support scalable, governance-compliant analysis of population-scale genomic datasets. The Nextflow–Lifebit architecture ensured reproducibility, provenance tracking, and fault-tolerant execution while conforming to GEL’s secure operational model. Even under financially constrained computational budgets, the workflow provided consistent and interpretable gene-level burden estimates suitable for downstream statistical modelling and machine-learning applications. Expanding future iterations of the workflow to encompass whole-genome non-coding regions, including enhancers, promoters, and regulatory domains, will be an important step towards fully characterising polygenic architecture and understanding the distributed pathogenicity landscape underlying complex traits. Complementing this will be the integration of additional deleteriousness metrics and functional priors tailored to non-coding annotations, which are expected to further enhance the interpretive power of gene-based burdens across the genome.

## Data Availability

All genomic data analysed in this study were accessed and processed within the Genomics England Research Environment via the Lifebit CloudOS platform. Data are available to approved researchers through the Genomics England Research Environment at https://www.genomicsengland.co.uk/research.
Analysis code and pipeline scripts are available at https://github.com/UoS-HGIG/Genepy_GEL_V2

## Acknowledgements

We gratefully acknowledge the participants of the National Genomic Research Library (NGRL), whose contributions made this research possible. Secure access to the NGRL under project ID [1055] was provided by Genomics England, which delivers the NGRL in partnership with NHS England and is wholly owned by the UK Department of Health and Social Care. The NGRL contains participants’ health data collected by the NHS as part of their care, along with samples and data from their participation in research, for which fully informed consent has been obtained. This includes genomic and clinical data provided through the NHS Genomic Medicine Service, as well as data obtained through research studies, including the 100,000 Genomes Project and the Generation Study, both of which are delivered in partnership with the NHS, and from other research cohorts involving external collaborators.

We also acknowledge the Lifebit engineering and support teams for their technical assistance and for maintaining the computational infrastructure used throughout this work. We are grateful to our colleagues within the research group for their guidance and continued support, and we thank all contributors and participants whose involvement made this research possible. This study was supported by the National Institute for Health Research (NIHR) Southampton Biomedical Research Centre. The views expressed are those of the author(s) and not necessarily those of the NIHR or the Department of Health and Social Care.

## Funding

JJA is funded by an NIHR Advanced Fellowship (NIHR302478).

## Conflict of interest

JJA is a scientific advisory board member for Orchard Therapeutics, he has contributed to a personalised IBD think tank (Takeda funded) and participated in a IBD transition Delphi consensus process (Pfizer funded). The other authors declare no conflicts of interest in relation to the submitted work.

## Data Access Statement

Data from the National Genomic Research Library (NGRL) used in this research are available within the secure Genomics England Research Environment. Access to NGRL data is restricted in order to adhere to consent requirements and protect participant privacy.

Access to NGRL data is provided to approved researchers who are members of the Genomics England Research Network, subject to institutional access agreements and research project approval under participant-led governance. For more information on data access, visit https://www.genomicsengland.co.uk/research.

## Notes

### Competing Interest Statement

The authors have declared no competing interest.

## References

[1] Mossotto E, Ashton JJ, O’Gorman L, Pengelly RJ, Beattie RM, MacArthur BD, Ennis S. GenePy - a score for estimating gene pathogenicity in individuals using next-generation sequencing data. BMC Bioinformatics. 2019;20(1):254. doi:10.1186/s12859-019-2877-3

[2] Seaby EG, Leggatt G, Cheng G, Thomas NS, Ashton JJ, Stafford I, et al. A gene pathogenicity tool “GenePy” identifies missed biallelic diagnoses in the 100,000 Genomes Project. Genetics in Medicine. 2024;26(4):101073. doi:10.1016/j.gim.2024.101073

[3] Genomics England. The National Genomic Research Library. 2024. Available at: 10.6084/m9.figshare.4530893

[4] Di Tommaso P, Chatzou M, Floden EW, Barja PP, Palumbo E, Notredame C. Nextflow enables reproducible computational workflows. Nature Biotechnology. 2017;35:316–319. doi:10.1038/nbt.3820

[5] Merkel D. Docker: lightweight Linux containers for consistent development and deployment. Linux Journal. 2014;2014(239):2.

[6] Kurtzer GM, Sochat V, Bauer MW. Singularity: Scientific containers for mobility of compute. PLoS ONE. 2017;12(5):e0177459. doi:10.1371/journal.pone.0177459

[7] Lifebit. Lifebit CloudOS data intelligence platform. Available at: https://lifebit.ai

[8] Amazon Web Services. Amazon Web Services cloud platform. Available at: https://aws.amazon.com

[9] McLaren W, Gil L, Hunt SE, et al. The Ensembl Variant Effect Predictor. Genome Biology. 2016;17:122. doi:10.1186/s13059-016-0974-4

[10] Chen S, Francioli LC, Goodrich JK, et al. A genomic mutational constraint map using variation in 76,156 human genomes. Nature. 2024;625:92–100. doi:10.1038/s41586-023-06045-0

[11] Rentzsch P, Schubach M, Shendure J, Kircher M. CADD-Splice—improving genome-wide variant effect prediction using deep learning-derived splice scores. Genome Medicine. 2021. doi:10.1186/s13073-021-00835-9

[12] Carson AR, Smith EN, Matsui H, et al. Effective filtering strategies to improve data quality from population-based whole exome sequencing studies. BMC Bioinformatics. 2014;15:125. doi:10.1186/1471-2105-15-125

[13] Kim S, Scheffler K, Halpern AL, et al. Strelka2: fast and accurate calling of germline and somatic variants. Nature Methods. 2018;15(8):591–594. doi:10.1038/s41592-018-0051-x

[14] Graffelman J, Morales-Camarena J. Graphical tests for Hardy–Weinberg equilibrium based on the ternary plot. Human Heredity. 2008;65(2):77–84. doi:10.1159/000108939

